# Weekly Intra-Treatment Diffusion Weighted Imaging Dataset for Head and Neck Cancer Patients Undergoing MR-linac Treatment

**DOI:** 10.1101/2023.08.18.23294280

**Authors:** Dina M El-Habashy, Kareem A Wahid, Renjie He, Brigid McDonald, Samuel J. Mulder, Yao Ding, Travis Salzillo, Stephen Lai, John Christodouleas, Alex Dresner, Jihong Wang, Mohamed A Naser, Clifton D Fuller, Abdallah Sherif Radwan Mohamed

## Abstract

Radiation therapy (RT) is a crucial treatment for head and neck squamous cell carcinoma (HNSCC), however it can have adverse effects on patients’ long-term function and quality of life. Biomarkers that can predict tumor response to RT are being explored to personalize treatment and improve outcomes. While tissue and blood biomarkers have limitations, imaging biomarkers derived from magnetic resonance imaging (MRI) offer detailed information. The integration of MRI and a linear accelerator in the MR-Linac system allows for MR-guided radiation therapy (MRgRT), offering precise visualization and treatment delivery. This data descriptor offers a valuable repository for weekly intra-treatment diffusion-weighted imaging (DWI) data obtained from head and neck cancer patients. By analyzing the sequential DWI changes and their correlation with treatment response, as well as oncological and survival outcomes, the study provides valuable insights into the clinical implications of DWI in HNSCC.

|  |  |
| --- | --- |
| Design Type(s) | Data integration objective |
| Measurement Type(s) | Primary tumor and nodal target volumes |
| Technology Type(s) | MRI scan (DWI) |

## 1. Background & Summary

Radiotherapy (RT) is a cornerstone of head and neck squamous cell carcinoma (HNSCC) treatment, both as a primary treatment option and as a post-operative therapy ^1^. While RT is effective in treating this type of cancer, it can also have adverse effects that may impact patient’s long-term function and quality of life ^2^. The response of tumors to RT varies, and predicting this response using a specific biomarker could help tailor RT doses and potentially improve treatment outcomes with reduced toxicity ^3^.

Various biomarkers derived from tissue or blood have been extensively studied to determine their role in guiding personalized clinical decisions in HNSCC ^4^. Among these biomarkers, only human papillomavirus (HPV) has recently accredited as a predictive and prognostic biomarker specially for oropharyngeal cancer ^5^. Despite their usefulness, tissue and blood biomarkers have certain limitations. Tissue markers provide information from a small region of the tumor, usually obtained at a single time point, resulting in limited spatial and temporal resolution. On the other hand, blood biomarkers, such as liquid biopsies, offer a comprehensive overview of the tumors’ secreted factors but lack spatial resolution entirely. In contrast, imaging biomarkers can evaluate each tumor volume, including primary tumors and lymph node metastases, individually. Furthermore, they can be obtained at multiple time points, thereby, offering superior temporal and spatial resolution compared to either tissue or blood biomarkers ^6^.

Magnetic resonance imaging (MRI) provides detailed anatomical and functional information regarding the tumor. The potential benefits of utilizing MRI in tumor delineation and assessing their response to RT has prompted international collaborations to develop MR-linac technology ^7^. MR-linac represents an innovative RT device that combines MRI and a linear accelerator, allowing for the acquisition of quantitative images on a daily basis. This integration establishes the basis of MR-guided radiation therapy (MRgRT), enabling precise and real-time visualization and treatment delivery ^8^.

Diffusion-weighted imaging (DWI) is an MRI technique with potential utility for assessing tumor response by providing functional information regarding the movement of water molecules into intra/inter-cellular spaces, which is largely affected by cellularity within tumors. The quantification of water diffusion in each voxel is assessed using the apparent diffusion coefficient (ADC), a quantitative imaging biomarker ^9^.

Multiple studies have investigated the potential of ADC as a biomarker in head and neck cancer ^10-15^. However, no studies in head and neck cancer have yet incorporated regular interval DWI scans throughout the course of RT using MR-linac device.

Herein, we aim to analyze the sequential quantitative changes in DWI within the primary tumor and lymph node metastases in patients with head and neck cancer who received RT using an MR-Linac device. This dataset offers a unique opportunity to leverage frequent DWI scans throughout the entire course of RT, enabling the quantification of weekly ADC changes (ΔADC) (Figure 1). Additionally, these changes could be correlated with RT response, as well as oncological and survival outcomes, providing valuable insights into clinical implications of DWI in head and neck cancer treatment.

**Figure 1.**
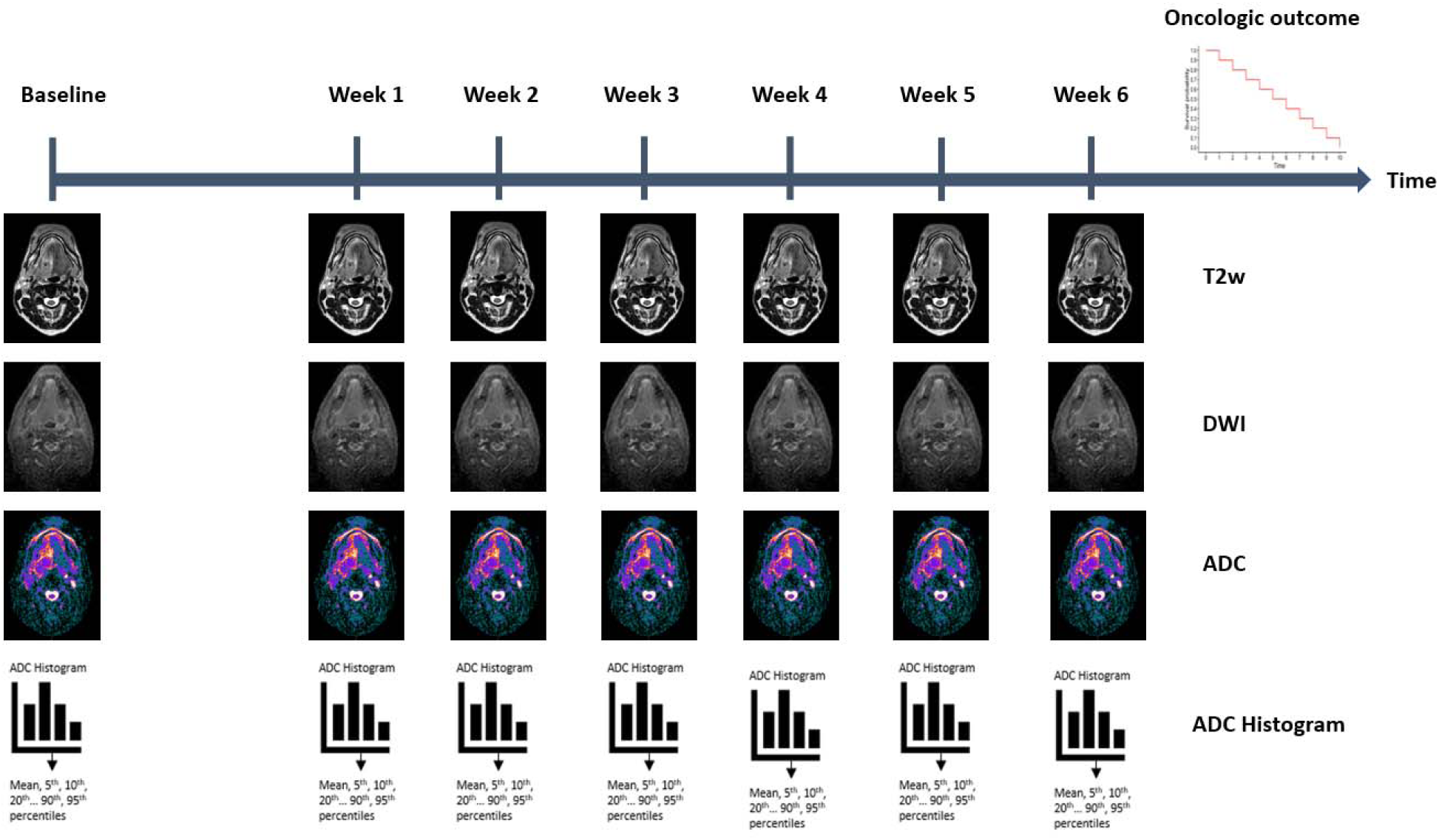
Overview of the study design. T2w: T2 weighted DWI: Diffusion weighted image ADC: Apparent diffusion coefficient

## 2. Methods

### 2.1 Patient Population

In this pilot study, 30 patients with HNSCC treated with curative-intent intensity modulated radiation therapy (IMRT) at The University of Texas MD Anderson Cancer Center between May 2019 to February 2021 were included. The enrolled patients were participants of the Multi-OutcoMe EvaluatioN of radiation Therapy Using the Unity MR-Linac Study (MOMENTUM), which is an observational registry conducted across multiple institutions. MOMENTUM focuses on patients who received treatment using the MR-linac system and is registered under the identifier NCT04075305.

Approval for the study was obtained from the Institutional Review Board (Protocol number: PA18-0341) at MD Anderson Cancer Center.

Patients included in this study should have fulfilled the following criteria:

1. Diagnosis of HNSCC is pathologically confirmed.
2. Curative-intent IMRT, with or without chemotherapy, was received using MR-linac device.
3. Patients with performance Status (PS) of 0-2 according to the ECOG (Eastern Cooperative Oncology Group) Score for Cancer Patients by Oncology Healthcare Professionals ^16^.
4. No contraindications for MRI ^17^.

The following data were gathered utilizing the EPIC electronic medical record system.

#### 2.1.1 Demographic data

The patients’ demographic data included: age at diagnosis, gender, ethnicity, and smoking status. Smoking status at diagnosis was documented as current smoker, former smoker and never-smoker, based on the 2023 ICD 10 definitions ^18^.

#### 2.1.2 Disease-related Data

The initial evaluation of the disease involved a comprehensive history and physical examination. Subsequently, nasopharyngolaryngoscopy was conducted to assess the site and extent of primary tumor with biopsies taken from suspicious areas. For better staging of HNSCC, all patients underwent contrast-enhanced CT (CECT), MRI and/or positron emission tomography-computed tomography (PET/CT) scans of the head and neck. Surgery was primarily implemented for diagnostic purposes, and was typically performed before RT.

Disease characteristics encompassed: tumor laterality, head and neck specific subsite of origin, tumor histology and grade, tumor stage, and HPV status. TNM (Tumor, node and metastases) classification was also provided according to the American Joint Committee on Caner (AJCC), 8^th^ edition ^19^.

Patients with squamous cell carcinoma (SCC) of the oropharynx, neck metastases without an identifiable primary tumor (Carcinoma of Unknown Primary), and laryngeal SCC in the absence of a smoking history were recommended to undergo HPV testing ^20^. Initially, p16 expression was confirmed using immunohistochemistry (IHC) for HPV detection. Subsequently, tumors that exhibited positive p16 expression underwent more rigorous HPV-specific detection methods, such as HPV DNA in situ hybridization (ISH) and/or a PCR-based assay. Conversely, tumors that were negative for p16 expression were considered HPV-unrelated and did not require further intervention ^21^. For the whole cohort, the HPV status was categorized as positive, negative or unknown.

#### 2.1.3 Treatment Data

Details of RT course were described and included:

- Total dose of irradiation each patient received in Grays.
- Dose per fraction received in Grays.
- Total number of daily radiation treatment fractions.
- Dates of start and end of RT.

Systemic treatment eligibility and the choice of treatment regimens were based on factors such as the disease extent, PS, and comorbidities. Therefore, patients with a significant tumor burden and/or large metastatic lymph nodes were commonly prescribed induction chemotherapy (i.e., before the initiation of radiation treatment course) and/or concurrent chemoradiation (i.e., simultaneously during the course of RT). The administration of systemic treatment (for both the induction phase and/ or the concurrent phase) was reported as a binary variable “0=yes or 1=no”.

### 2.2 Details of Treatment Technique

Initially, simulation was conducted using a non-contrast CT scan, which was obtained for the patient while in the supine-neck extended position. External room lasers and a scout film were used for patient alignment. A mouth opening/ tongue depressing stent was inserted to aid in positioning and immobilization. Custom thermoplastic masks, a posterior head cradle, and non-custom shoulder pulls were utilized to ensure reproducible set-up for radiation treatments. Isocenter for treatment planning was placed superior to arytenoids, and CT images from the vertex to carina were obtained.

Subsequently, the patient was brought to the MR-linac (Elekta AB, Stockholm, Sweden) and placed in the treatment position using the previously mentioned immobilization devices from the initial simulation. Axial MRIs were obtained for the region of interest, and these images were then transferred to the Monaco 5.4 (Elekta AB, Stockholm, Sweden) treatment planning system (TPS).

IMRT was delivered using the Elekta Unity system, which combines a 1.5T MRI system and a gantry positioned around the isocenter. The gantry houses a 7 MV linear accelerator with a flattening filter free (FFF) configuration ^22^. Blanchard at al. provided a detailed description of the schematic IMRT that was administrated to all patients ^23^.

### 2.3 Clinical Outcomes

Response to treatment was categorized as LJ0=occurrence of complete remission (CR)” or LJ1=no CR” based on the Response Evaluation Criteria in Solid Tumors (RECIST) guidelines, version 1.1 ^24^. The assessment of treatment response was conducted either weekly during RT, using the MR images obtained from the MR-linac system, or post-RT completion.

Confirmation of recurrence required pathological analysis and was denoted as LJ0= tumor recurrence” or LJ1=no tumor recurrence”. Recurrence was further subcategorized as LJlocal” if it occurred within the same subsite of the primary tumors, LJregional” if it occurred in the neck, or LJdistant” if it occurred outside the head and neck region. In the same context, patient’s vital status was reported as a binary outcome LJ0=alive” or LJ1=dead”; as an indicator for overall survival status.

### 2.4 Image Details

MRI scans were obtained using the Unity system (Elekta AB, Stockholm, Sweden), based on a Philips 1.5 T Marlin MRI equipped with a 4-element anterior coil and a built-in 4 element posterior coil, providing comprehensive coverage of the head and neck region. The MRIs were acquired at baseline (Pre-RT) and weekly at a regular interval throughout the entire 7-week RT course. The following images were utilized in the study:

#### 2.4.1 T2 weighted images

1. Three-dimensional (3D) T2-weighted MRI (T2 3D Tra):

- These were utilized for image registration during the daily treatment process (slice thickness = 2, repetition time = 1535 ms, echo time = 278 ms, imaging frequency = 64, spacing between slices = 1, number of phase encoding steps = 268, percentage phase field of view = 100, pixel bandwidth = 740 Hz, flip angle = 90°, echo train length = 114, reconstruction diameter = 400, field of view = 400×400×300 mm^3^, resolution = 1.2000 pixels/mm, reconstructed voxel size = 0.83×0.83×1 mm^3^, scan time = 2 minutes, number of average = 1, and SENSE factor = 4)
- Total number of scans = 172.
2. Three-dimensional T2-weighted MRI without fat suppression (T2 3D Tra):

- These MRI sequences were used for tumor segmentation purposes (slice thickness = 2.2, repetition time = 2100 ms, echo time = 375 ms, imaging frequency = 64, spacing between slices = 1.1, number of phase encoding steps = 433, percentage phase field of view = 57, pixel bandwidth = 459 Hz, flip angle = 90°, echo train length = 150, reconstruction diameter = 520, field of view = 520×520×300 mm^3^, resolution = 1.2000 pixels/mm, reconstructed voxel size = 0.98×0.98×2.2 mm^3^, scan time = 6 minutes, number of average = 2, and SENSE factor = 2)
- Total number of scans = 9.
3. Three-dimensional T2-weighted MRI with fat suppression (3D T2 SPAIR):

- Used for target segmentation purposes (slice thickness = 2, repetition time = 1400 ms, echo time = 190 ms, imaging frequency = 64, spacing between slices = 1, number of phase encoding steps = 345, percentage phase field of view = 52, pixel bandwidth = 473 Hz, flip angle = 90°, echo train length = 76, reconstruction diameter = 520, field of view = 520×520×300 mm^3^, resolution = 1.2308 pixels/mm, reconstructed voxel size = 0.98×0.98×1.2 mm^3^, scan time = 6 minutes, fat saturation = SPAIR, number of average = 2, and SENSE factor = 2)
- Total number of scans = 1.

#### 2.4.2 DWI

Single-shot echo planar DWI was utilized for evaluation of treatment response (b values = 0, 150, and 500 s/mm^2^, slice thickness = 4, repetition time = 5700 ms, echo time = 75 ms, inversion time = 180, imaging frequency = 64, spacing between slices = 4, number of phase encoding steps = 86, percentage phase field of view = 100, pixel bandwidth = 2174 Hz; flip angle = 90°, echo train length = 39, reconstruction diameter = 300, field of view = 300×300×158 mm^2^, resolution = 0.64 pixels/mm, reconstructed voxel size = 1.6×1.6×1.3 mm^3^; scan time = 3 minutes, fat saturation = SPAIR, and SENSE factor = 2.2).

It is noteworthy that we followed the consensus EPI protocol provided by the MR-linac Consortium ^25^.

#### 2.4.3 ADC maps

ADC maps were created using b-values of 150 and 500 s/mm², with the exclusion of b=0 images. This approach was chosen to reduce the impact of perfusion on ADC calculations and to align with the guidelines set forth by the MR-linac Consortium ^26^.

The imaging data were presented in the standardized format of Digital Imaging and Communications in Medicine (DICOM).

### 2.5 ADC Parameter Calculation

An in-house MATLAB script (MATLAB, MathWorks, MA, USA) was employed to process the ADC maps, extracting histogram parameters (mean, 5^th^, 10^th^, 20^th^, 30^th^, 40^th^, 50^th^, 60^th^, 70^th^, 80^th^, 90^th^, and 95^th^ percentiles) for the segmented regions of interest (ROIs).

The ADC was commonly calculated using DWI data with at least two non-zero b-values (b values = 0, 150, and 500 s/mm^2^). However, since the b0 image (non-diffusion-weighted image) is affected by the perfusion phenomenon, an alternative method is used to calculate the ADC parameter. The following is a general overview of the approach:

1. Acquire DWI data: Obtain multiple DWIs with different non-zero b-values.
2. Preprocessing: Apply necessary preprocessing steps to correct for artifacts, distortions, and noise reduction.
3. ADC calculation: The ADC can be calculated without the b0 image using the following equation: ADC = ln(S1/S2) / (b1 - b2) where S1 and S2 are the signal intensities of two diffusion-weighted images with different b-values (b1 and b2, respectively). The natural logarithm (ln) is applied to the ratio of the signal intensities, and the result is divided by the difference in b-values.
4. ADC map generation: Apply the calculated ADC values to create an ADC map, where each pixel represents the ADC value.
5. Data interpretation: Analyze the ADC map to assess tissue characteristics. Lower ADC values indicate restricted diffusion, which may be associated with increased cellularity or tissue pathology, while higher ADC values suggest increased diffusion and decreased tissue density. The appropriate software and techniques specific to our research and clinical setting is based on above principles for accurate and reliable ADC parameter calculation.

### 2.6 Target Volume Segmentation

The primary disease and nodal volumes segmented on the baseline MRI were named as DGTVP-BL and GTV-N”, respectively. Segmentation was conducted on each weekly MRI for both GTV-N and the residual primary tumor disease volume, referred to as DGTVP-RD”. Additionally, responding sub-volumes of the primary tumors were generated by subtracting GTVP-RD from GTVP-BL and labeled as DGTVP-RS”. A visual representation of the various target volumes is illustrated in Figure 2. Segmented structures were saved in DICOM Radiotherapy Structure (DICOM RTS) format.

**Figure 2.**
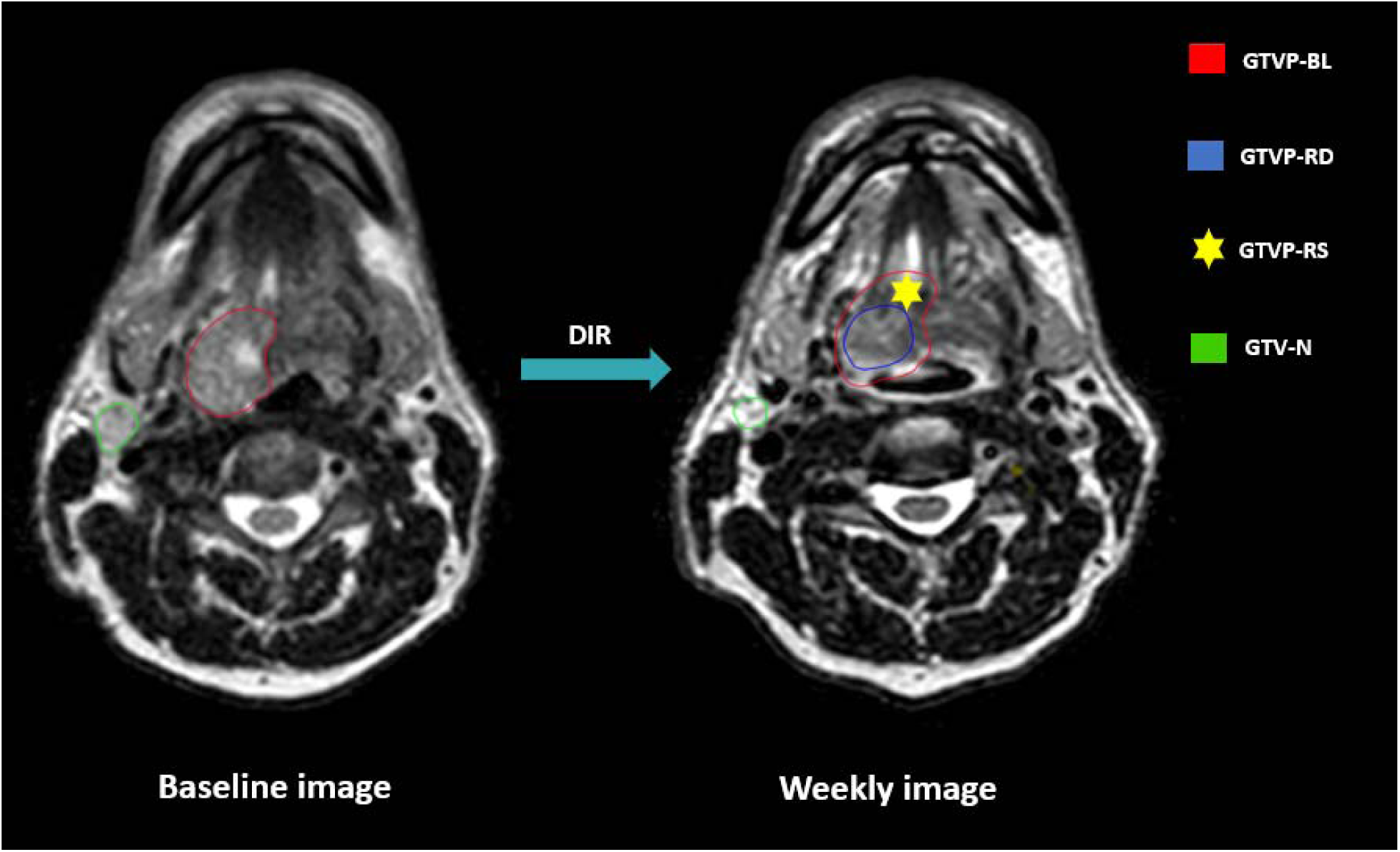
Gross Tumor volume segmentation. GTVP-BL: Baseline primary tumor volume GTVP-RD: The residual primary disease volume GTVP-RS: The volume of the primary disease responding to RT GTV-N: Gross nodal disease volumes DIR: Deformable image registration

### 2.7 Image Registration

Baseline T2-weighted images were utilized for manual segmentation of baseline GTV-P and GTV-N, respectively. Different ROIs were propagated to the rigidly co-registered DWIs at the same timepoint. Afterward, deformable image registration (DIR) was conducted to align the images from different weeks with the baseline images. Both manual segmentation and registration processes were executed using Velocity AI 3.0.1 software (Atlanta, GA, USA).

## 3. ADC Parameter Data Imputation

To address the issue of missing ADC parameter values resulting from a few patients having incomplete weekly image data, imputation was performed using an order-1 linear spline method, i.e., a piecewise linear function used to interpolate between given data points. An order-1 linear spline was chosen due to the relationship between ADC parameters and time not being strictly monotonic (e.g., parameters could increase in value initially and then decrease in value). Specifically, for each ROI for each patient, timepoint labels were converted to an integer scale as follows: pre-RT = 0, weeks 1-6 = 1-6, post-RT = 7. Parameters were then imputed based on the availability of existing timepoint values. As a hypothetical example, if parameter values of only 1, 5, 8, and 2 were available for timepoints 0, 3, 4, and 7, a set of linearly increasing values between 1 and 5 would be imputed for timepoints 1 and 2 while a set of linearly decreasing values between 8 and 2 would be imputed for timepoints 5 and 6. Since the imputation could yield negative values, but ADC parameters could only be positive, all imputed values were truncated to 0. Imputation was only performed if at least 2 timepoints were available. This imputation process was carried out using an in-house Python script (Python version 3.8.8) utilizing the Pandas dataframe interpolate method using the following parameters: method = “slinear”, fill_value = “extrapolate”, limit_direction = “both”. As an example, for one parameter value, the imputed median ADC of GTVP-BL structures is shown in Figure 3.

**Figure 3.**
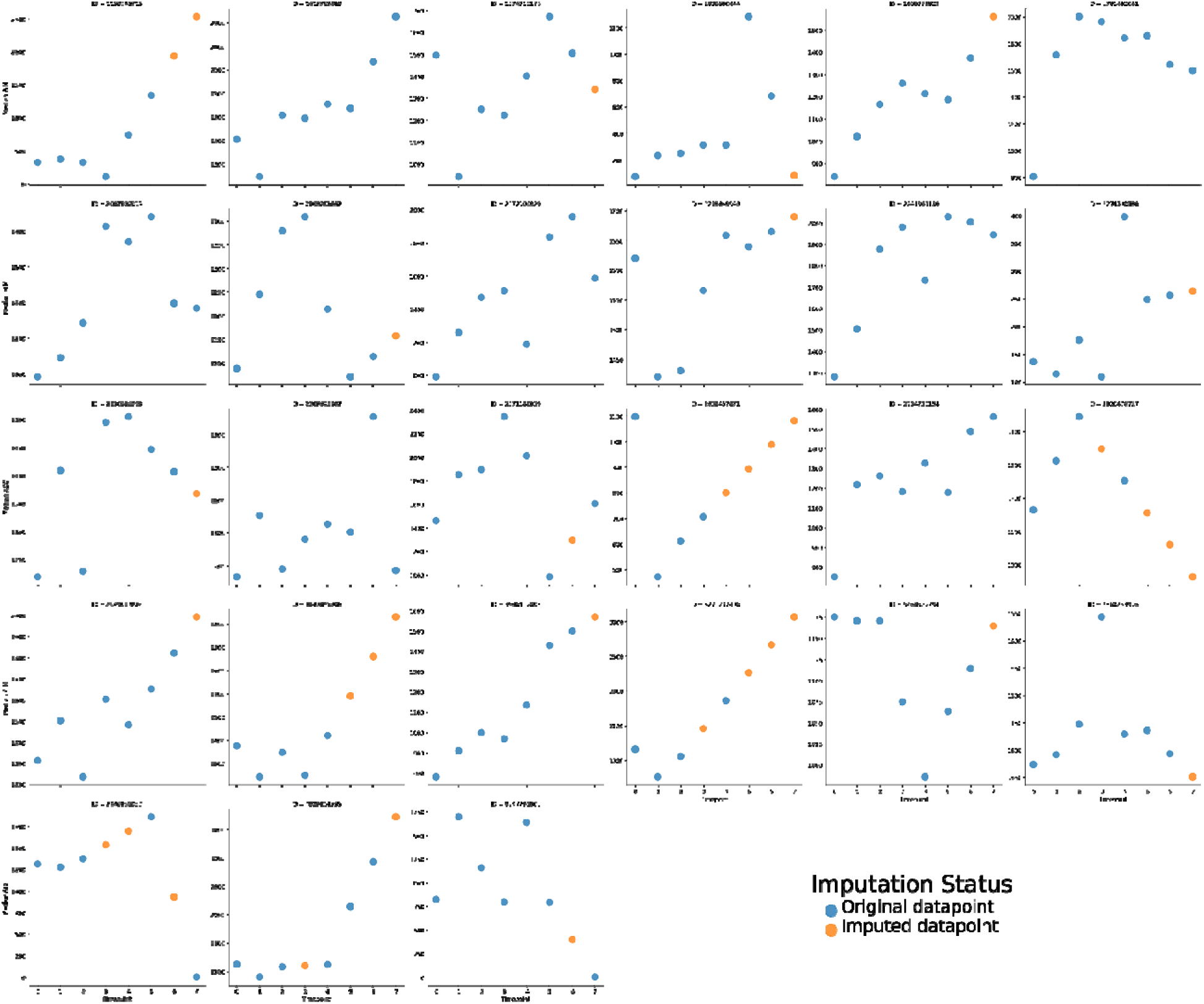
Baseline primary tumor imputed median ADC values for. Missing values were imputed using a linear spline method. Each subplot corresponds to a different patient, where different colored dots correspond to imputation status (original datapoint - blue, imputed datapoint - orange).

## 4. Data De-identification

DICOM data (images and RTS files) were anonymized using an in-house Python script that implements the RSNA CRP DICOM Anonymizer software. All files have had any DICOM header info and metadata containing PHI removed or replaced with dummy entries. Notably, patient medical record numbers were mapped to new randomized numeric values, which serve as their anonymized identifiers, and any corresponding date data were mapped to new randomized dates which preserved the relative order and time between the image acquisitions.

## 5. Structure Name Cleaning

Due to the presence of misspellings or duplicate structures in exported DICOM RTS structure names, structures names were harmonized using the Pydicom v. 2.2.2 Python library ^27^.

## 6. Data Record

### 6.1 Segmentation Data (Structures & ADC maps)

In total, 7537 anonymized DICOM ADC map image files and 200 anonymized RTS files are provided for this dataset. Figure 4 and Figure 5 demonstrate the total number of images and structures included in this dataset when stratified by timepoint, respectively. The folder structure for the anonymized dataset is shown in Figure 6.

**Figure 4.**
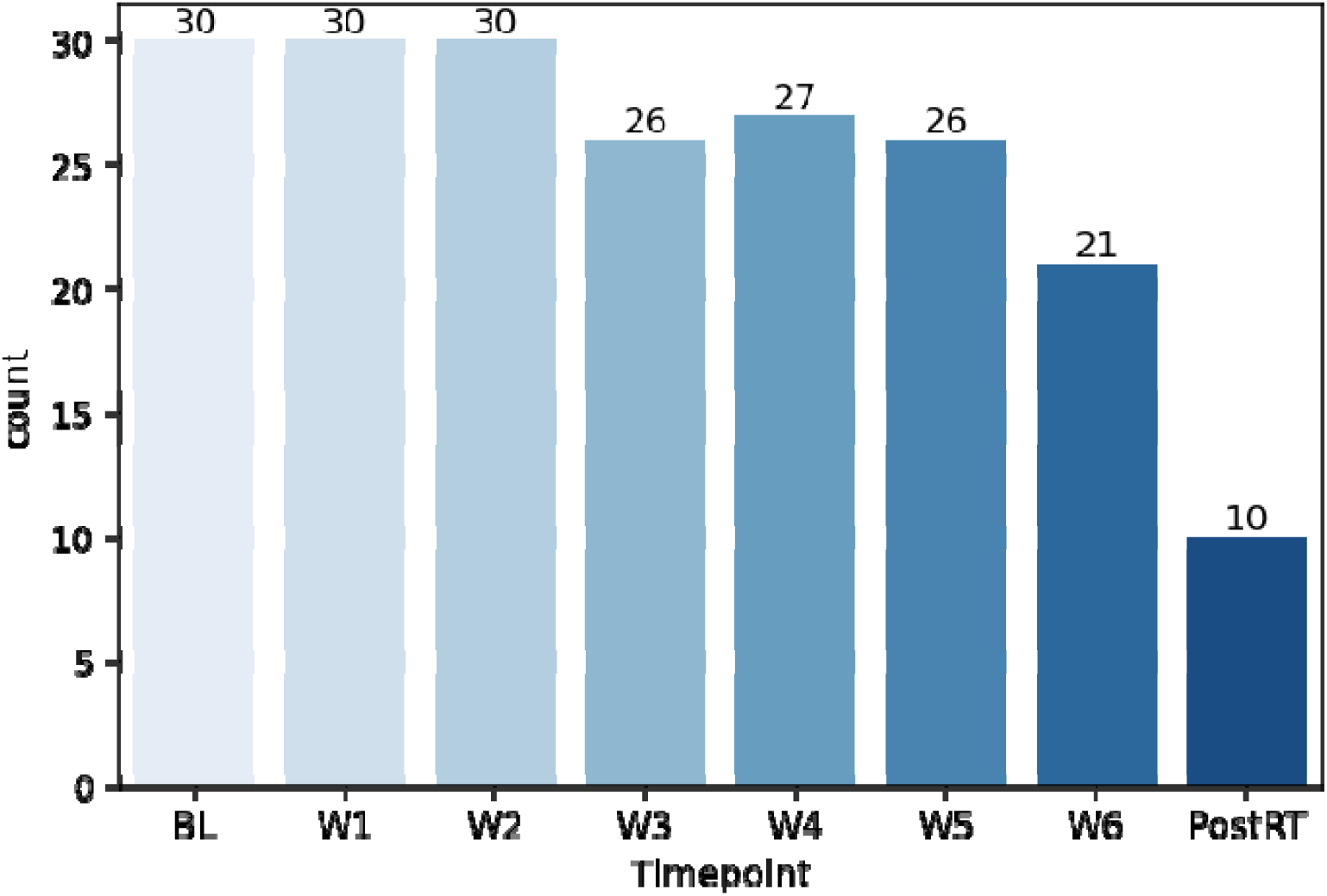
Number of images available at each timepoint.

**Figure 5.**
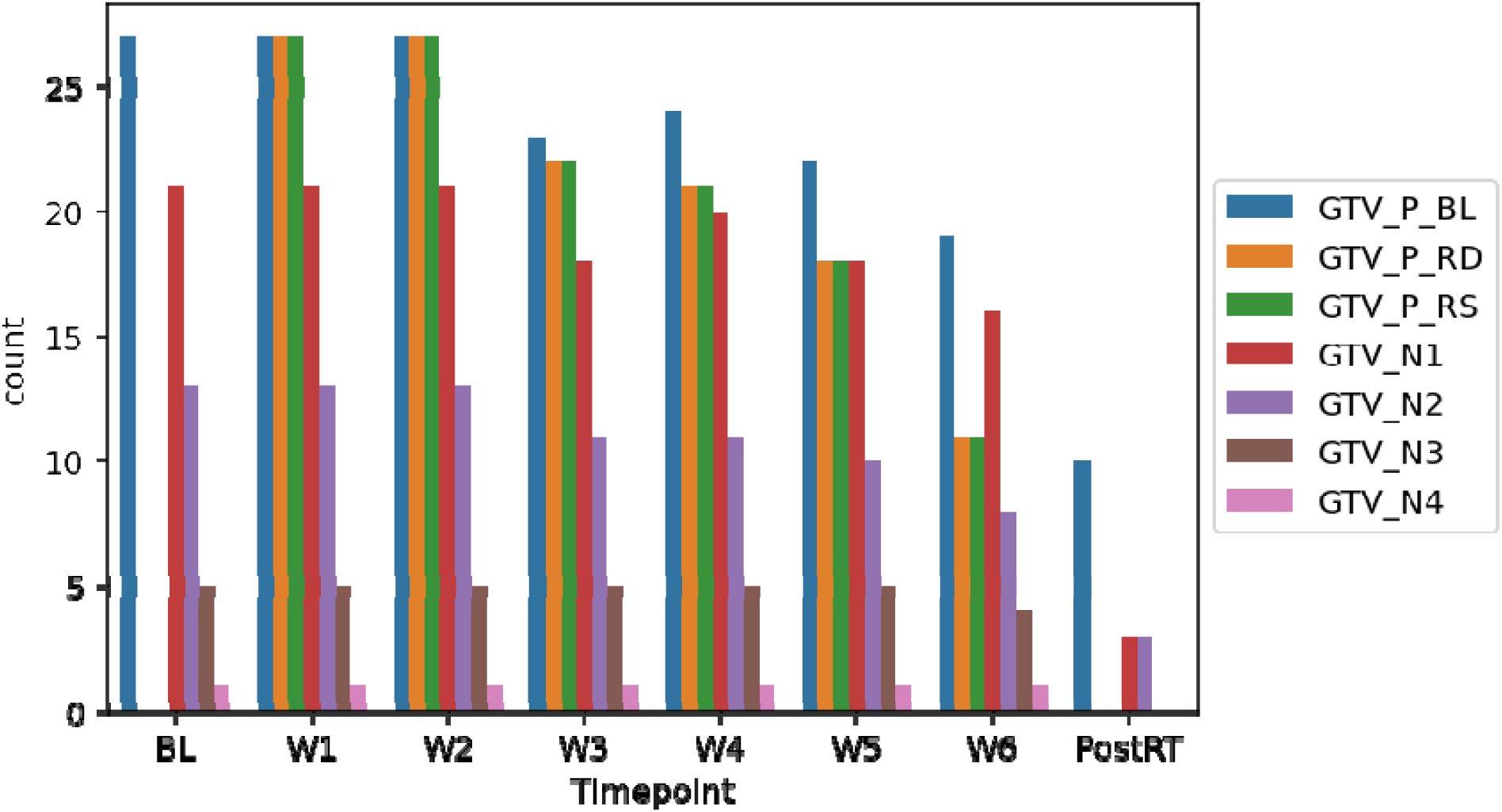
Number of structures available at each timepoint.

**Figure 6.**
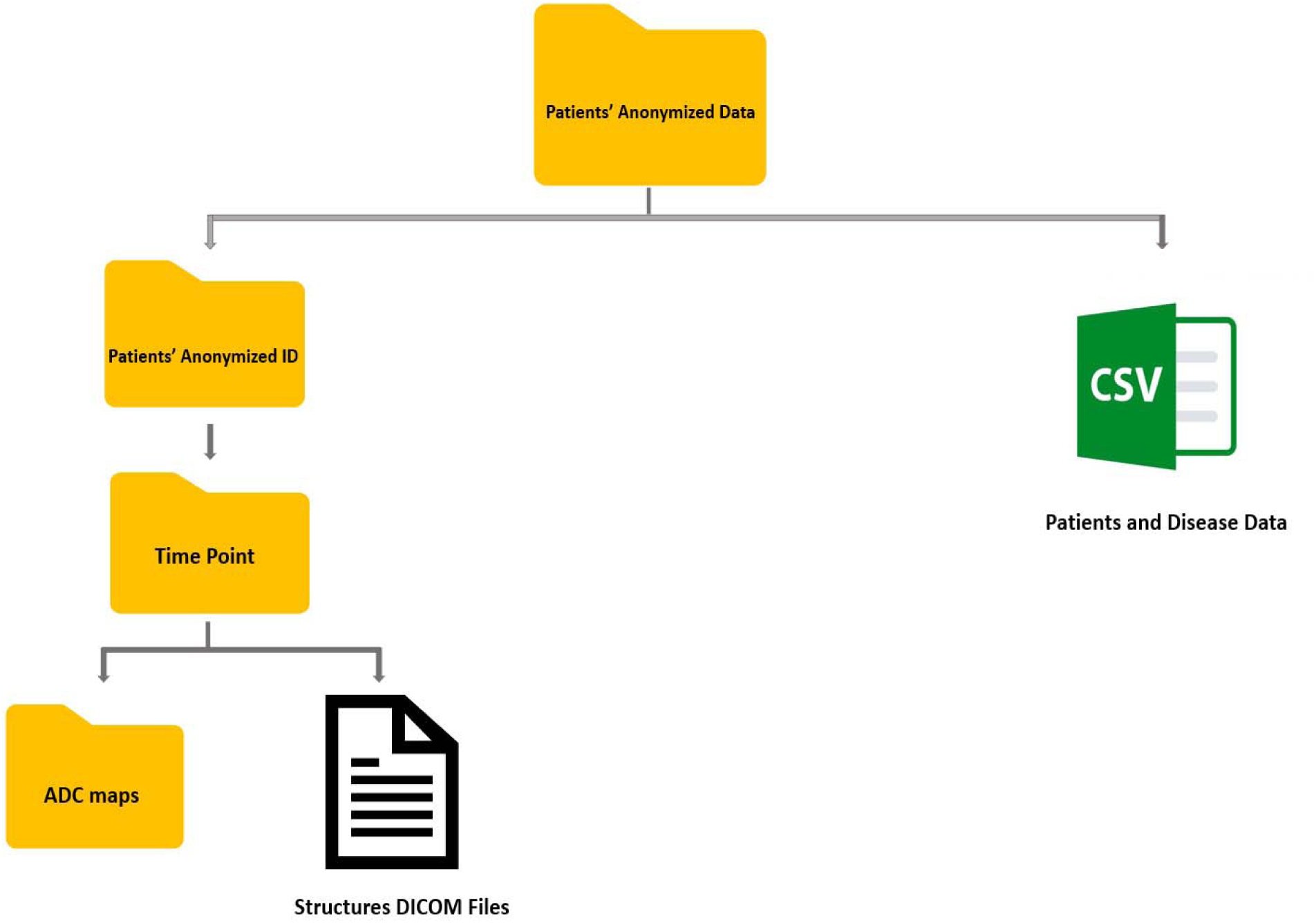
The anonymized dataset folder structure.

### 6.2 Clinical Data

We offer a comprehensive comma-separated value (CSV) file that includes the clinical, pathological and demographic data. By incorporating these data into the CSV file, we provide a consolidated resource for exploring the relationship between clinical variables, ADC measurements, and the response of head and neck cancer to treatment. The anonymized ADC maps, segmented structures, and clinical data are cited under Figshare; doi: 10.6084/m9.figshare.22766783.

## 7. Technical Validation

### 7.1 Segmentation

Target volumes were segmented and reviewed by two trained radiation oncologists: DE and ASRM, possessing 9 and 15 years of experience, respectively.

### 7.2 MR-linac

The technical validation of quantitative images acquired from the 1.5 T MR-Linac device serves as a critical process to establish the reliability and accuracy of these images as biomarkers. This validation process is essential to ensure that the acquired quantitative data can be consistently utilized as reliable indicators or measurements for specific biological characteristics or processes. Several studies have been conducted for this specific purpose ^28-30^.

### 7.3 EPIC (Electronic Medical Record System)

Clinical data (patients and disease characteristics) were manually collected from the University of Texas MD Anderson Cancer Center clinical databases through the EPIC electronic medical record system.

Data collection was conducted by a trained physician (DE). Epic is a well-known EHR software platform (https://www.clinfowiki.org/wiki/index.php/Epic_Systems).

## 8. Usage Notes

This data (Images and segmentations) is provided in DICOM format with the accompanying CSV file containing additional clinical information. We invite all interested researchers to download this dataset to use in researches about DWI kinetics analysis in cancer patients receiving RT.

## Data Availability

Data will be available under Figshare; doi: 10.6084/m9.figshare.22766783.

https://figshare.com/account/items/22766783/edit

## 9. Code Availability

Codes used for data annotation is available through GitHub: (https://github.com/kwahid/Weekly_DWI_Data_Descriptor)

## Authors Contributions

**Dina M El-Habashy:** Methodology, Writing - Original Draft, Writing - Review and Editing

**Kareem A Wahid**: Software, Data Curation, Writing - Review and Editing

**Renjie He**: Software, Writing - Review and Editing

**Brigid McDonald**: Writing - Review and Editing

**Samuel J. Mulder**: Writing - Review and Editing

**Jinzhong Yang**: Writing - Review and Editing

**Yao Ding**: Methodology

**Mohamed A Naser**: Data Curation

**Travis C Salzillo**: Writing - Review and Editing

**Stephen Y. Lai:** Writing - Review and Editing

**Alex Dresner:** Writing - Review and Editing

**John Christodouleas:** Funding Acquisition, Writing - Review and Editing

**Abdallah Sherif Radwan Mohamed:** Funding Acquisition, Supervision, Writing - Review and Editing

**Clifton D Fuller:** Funding Acquisition, Supervision, Writing - Review and Editing

